# Automated Detection of Candidate Subjects with Cerebral Microbleeds using Machine Learning

**DOI:** 10.1101/2021.09.21.21263298

**Authors:** Vaanathi Sundaresan, Christoph Arthofer, Giovanna Zamboni, Robert A. Dineen, Peter M. Rothwell, Stamatios N. Sotiropoulos, Dorothee P. Auer, Daniel Tozer, Hugh S. Markus, Karla L. Miller, Iulius Dragonu, Nikola Sprigg, Fidel Alfaro-Almagro, Mark Jenkinson, Ludovica Griffanti

## Abstract

Cerebral microbleeds (CMBs) appear as small, circular, well defined hypointense lesions of a few mm in size on T2*-weighted gradient recalled echo (T2*-GRE) images and appear enhanced on susceptibility weighted images (SWI). Due to their small size, contrast variations and other mimics (e.g. blood vessels), CMBs are highly challenging to detect automatically. In large datasets (e.g. the UK Biobank dataset), exhaustively labelling CMBs manually is difficult and time consuming. Hence it would be useful to preselect candidate CMB subjects in order to focus on those for manual labelling, which is essential for training and testing automated CMB detection tools on these datasets. In this work, we aim to detect CMB candidate subjects from a larger dataset, UK Biobank, using a machine learning-based, computationally light pipeline. For our evaluation, we used 3 different datasets, with different intensity characteristics, acquired with different scanners. They include the UK Biobank dataset and two clinical datasets with different pathological conditions. We developed and evaluated our pipelines on different types of images, consisting of SWI or GRE images. We also used the UK Biobank dataset to compare our approach with alternative CMB preselection methods using non-imaging factors and/or imaging data. Finally, we evaluated the pipeline’s generalisability across datasets. Our method provided subject-level detection accuracy *>* 80% on all the datasets (within-dataset results), and showed good generalisability across datasets, providing a consistent accuracy of over 80%, even when evaluated across different modalities.

## 1. Introduction

Cerebral microbleeds (CMBs) represent focal haemosiderin depositions consisting of macrophages in microhaemorraghes (Shoamanesh et al., 2011), and are sometimes surrounded by ischemic areas and gliosis (Gouw et al., 2011). CMBs appear as small focal dark circular lesions on T2*-weighted gradient recalled echo (T2*-GRE) sequences. They usually range from 2 to 10 mm in size, although further subdivision into microbleeds (2-5 mm) and macrobleeds (*>*5mm) has been used (Greenberg et al., 2009a). While some of the CMBs might not be visible at all on T2-GRE images, susceptibility weighted images (SWI) shows more CMBs and they appear more prominently on SWI due to the blooming effect (Greenberg et al., 2009b; Charidimou and Werring, 2011). CMBs can be the early sign of intracerebral haemorrhage (ICH) (Gouw et al., 2011), vascular dementia (Ayaz et al., 2010) and Alzheimer’s disease (Gouw et al., 2011; Wardlaw et al., 2013; Shoamanesh et al., 2011). They have been associated with cognitive decline (Werring et al., 2004; Won Seo et al., 2007) and several vascular diseases (Nishikawa et al., 2009; Yates et al., 2014), including lacunar stroke and small vessel diseases (Nannoni et al., 2021). Moreover, CMBs often occur with vascular damage related to cerebral amyloid angiopathy (Gouw et al., 2011). Strong associations have been established between CMBs and risk factors such as age (Roob et al., 2000; Horita et al., 2003; Jeerakathil et al., 2004; Vernooij et al., 2008; Poels et al., 2010; Takashima et al., 2011), hypertension (Roob et al., 2000; Tsushima et al., 2002; Horita et al., 2003; Vernooij et al., 2008; Poels et al., 2010) and white matter damage (Roob et al., 2000).

Recent studies on the predictive value of CMBs for long-term cognitive outcome have shown inconsistent results, therefore the specific role of CMBs in cognitive impairment and neurodegeneration remains unclear. Hence, it would be useful to observe the prevalence and clinical/demographic associations of CMBs in larger populations. However, exhaustive manual labelling of CMBs is difficult and time consuming, especially in large datasets (e.g. the UK Biobank dataset). Consequently, semi-automated methods (Seghier et al., 2011; Barnes et al., 2011; van den Heuvel et al., 2016; Morrison et al., 2018; De Bresser et al., 2013; Kuijf et al., 2012, 2013) were proposed as a possible solution to reduce false positives. Even though the manual revising step removes the spurious detections effectively, it typically takes at least a few minutes per subject (van den Heuvel et al., 2016; Kuijf et al., 2013; Morrison et al., 2018) and it is not an efficient solution for CMB detection in very large datasets. Hence, it would be highly useful and efficient to develop a fully automated CMB candidate subject preselection method (without involving manual intervention in any stage of the detection pipeline) in the large datasets, and focus on those subjects for manual labelling to facilitate further semi-automated or fully-automated methods with more accurate CMB detection, analysis and characterisation (based on size, shape, location and multiplicity, clustering).

The detection of CMBs, however, even at subject-level, is highly challenging since CMBs occur sparsely, are difficult to detect due to their size, contrast variations and the fact that they are often accompanied by other signs (e.g. haemorrhages). The SWI modality has been shown to aid in identifying more CMBs (at least *>*67% (Nandigam et al., 2009)) compared to T2*-GRE images, since SWI improves CMB contrast. However, the presence of other paramagnetic substances, apart from haemosiderin, causes enhanced appearance of dark structures that resemble CMBs, known as CMB ‘mimics’ (Greenberg et al., 2009b; Charidimou and Werring, 2011). The mimics could be haemorrhagic/paramagnetic such as cavernous malformations, haemorrhagic micrometastases, diffusion axonal injury, small haemorrhages nearer to the infarcts and ICH areas, or non-haemorrhagic such as flow voids, calcifications, motion artefacts, Gibb’s ringing artefact and partial volume artefacts at air-bone interfaces (Greenberg et al., 2009b). For description and features of CMB mimics refer to suppl. table S1.

So far, the proposed automated methods for lesion-level CMB detection used shape descriptors (Bian et al., 2013; Fazlollahi et al., 2014, 2015), intensity and geometric information (Ghafaryasl et al., 2012), and location-based features (Dou et al., 2015). The use of comprehensive features, integrating the geometry, intensity, scale and local image structures from multiple modalities have been shown to improve CMB detection (Ghafaryasl et al., 2012; Dou et al., 2015). Over recent years, with the advent of deep learning, methods using convolution neural networks (CNNs) (Dou et al., 2016; Liu et al., 2019; Chen et al., 2018) and hybrid methods using a combination of CNNs and intensity information (Chen et al., 2015; Dou et al., 2015) have been proposed, occasionally with additional postprocessing steps (Liu et al., 2019). While CNN-based methods provide more accurate CMB detection when compared to conventional methods, they require a large amount of labelled training data. Alternatively, techniques such as semi-supervised and omni-supervised learning (Huang et al., 2018) requires more representative labelled CMB instances that would not bias the method towards CMB mimics. While the occurrence of CMBs could be high in disease groups (e.g. small vessel disease), in general, the prevalence of CMBs is low in population-based cohorts (e.g. UK Biobank). In datasets from populations with low prevalence of CMBs, it would be extremely time consuming to look at all subjects manually. A way to automatically preselect a subset that was enriched for CMBs would allow better use of the available manual identification time and lead to better and clinically relevant training datasets for further CMB analyses.

In this work, our aim is to develop a subject-level preselection method that is computationally light, easy to train and scalable to large datasets. Towards this aim, we propose a fully automated method using intensity, shape and location-based features for detecting CMB candidate subjects from large datasets such as the UK Biobank. We evaluated the method on datasets with different image modalities (GRE and SWI) and in the presence of other pathological signs. We then applied the method to the UK Biobank dataset for CMB candidate subject preselection, and compared our method with various alternative methods using non-imaging factors and/or imaging data (CMB count). We finally evaluated the ability of our proposed pipeline to adapt to differences in image characteristics and demographics of the datasets, by training our pipeline on one dataset and testing it on a different one.

## 2. Dataset details

In this work we used the following datasets to develop and evaluate the preselection pipeline. The datasets are diverse in their intensity characteristics and are acquired using different protocols. A brief overview of the datasets is provided below.

### The Oxford Vascular Study (OXVASC) dataset

The dataset consists of T2*-GRE images from 40 participants from the OXVASC study (Rothwell et al., 2004), who had recently experienced a minor non-disabling stroke or transient ischemic attack. The age range was 35.6 - 94.8 years, mean age 68.7 *±* 15.5 years, median age 67.4 years, and female to male ratio F:M = 15:25. The 2D single-echo T2*-GRE images were acquired using a 3T Siemens Verio scanner with GRAPPA factor = 2, TR/TE = 504/15 ms, flip angle 20^*o*^, voxel resolution of 0.9 × 0.8 × 5 mm, with image dimensions of 640 × 640 × 25 voxels. Out of 40 subjects, 20 subjects have CMBs and the corresponding manual segmentations, labelled on T2*-GRE images, are available. The total number of CMBs is 267, mean: 13.3 *±* 1.13 CMBs/subject.

### The Tranexamic acid for IntraCerebral Haemorrhage 2 (TICH2) trial MRI substudy dataset

This is a subset of the MRI dataset used in Dineen et al. (2018) obtained as part of the TICH2 trial (Dineen et al., 2018; Sprigg et al., 2018). The age range was 29 - 88 years, mean age 64.76 *±* 15.5 years, median age 66.5 years, and female to male ratio F:M = 24:26. The dataset consists of images acquired at multiple centres and on multiple MRI platforms with variations in image dimensions and voxel resolutions. MR acquisition parameters for the TICH2 MRI substudy dataset can be found in Dineen et al. (2018). The dataset used in this work consists of 50 SWI images from subjects with spontaneous intracerebral haemorrhage. Out of 50 subjects, 25 subjects have CMBs and manual segmentations for CMBs are available for all 25 subjects. The manual segmentations were labelled on SWI images as either definite or possible according to the microbleed anatomical rating scale (MARS) (Gregoire et al., 2009). Total number of CMBs: 505, mean: 20 *±* 32.6 CMBs/subject.

### UK Biobank (UKBB) dataset

Out of *≈* 20,000 subjects from the January 2018 release of UKBB, 14,521 had the required data fields (e.g. availability of SWI images and the factors specified below). From these subjects, we randomly selected 180 subjects with age range 46.8 - 76.8 years, mean age 58.9 *±* 9.1 years, median age 58.8 years, and female to male ratio F:M = 86:94. We used SWI images from the selected subjects for our experiments, which were constructed from 3D multi-echo GRE images acquired using a 3T Siemens Magnetom Skyra syngo MR D13 scanner with TR/TE1/TE2 = 27/9.4/20 ms, flip angle 15^*o*^, voxel resolution of 0.8 × 0.8 × 3 mm, with image dimensions of 256 × 288 × 48 voxels. Subject-level manual labels were available for the selected subjects, indicating whether each subject was a CMB or a non-CMB subject (only including those graded as definite CMB based on MARS scale). However, lesion-level manual segmentations of CMBs were not available for the dataset. On the UKBB dataset, we also used several non-imaging factors that are known to be associated with CMBs (given these data were collected as a part of the study), including demographic factors such as age (Charidimou and Werring, 2011; Greenberg et al., 2009b), blood pressure (BP) (Roob et al., 2000; Tsushima et al., 2002; Poels et al., 2010; Vernooij et al., 2008), risk factors such as smoking (Tsushima et al., 2002; Poels et al., 2010), clinical conditions such as white matter damage (Roob et al., 2000) and cognitive decline (Werring et al., 2004; Won Seo et al., 2007). We used the non-imaging factors for a comparison experiment on the dataset (for more details, refer to section 3.4.2). Summary statistics for the shortlisted factors for 180 subjects are: age (range provided above), BP (102/50 - 202/118 mmHg, mean: 134.7/80.7 *±* 42.4/25.3 mmHg, median: 147.5/90.5 mmHg), smoking (smoking:non-smoking = 70:110), white matter hyperintensity (WMH) volume (535 - 45,186 mm^3^, mean: 5,465 *±* 4,349 mm^3^, median: 2,929 mm^3^) and mean reaction time (MRT, as an indicator of cognitive ability) (397 - 896 ms, mean: 563 *±* 96.5 ms, median: 546 ms).

### 2.1. Data preprocessing

We reoriented the T2*-GRE and SWI images to match the orientation of the standard MNI template, retaining their original resolutions, in order to aid in further processing. We skull stripped the images using FSL BET (Smith, 2002), followed by bias field correction using FSL FAST (Zhang et al., 2001).

## 3. Pipeline for detection of CMB candidate subjects

The automated pipeline for CMB candidate subject preselection takes T2*-GRE images or SWI as input and provides a subject-level decision on whether the subject has CMBs or not. The pipeline consists of three steps: 1. removal of blood vessels and sulci, 2. voxel-wise detection of initial CMB candidates, 3. filtering of initial candidates using shape-based attributes.

### 3.1. Removal of blood vessels and sulci

In the first step, we removed the blood vessels, sulci and other elongated dark structures in the input image to reduce the prevalence of CMB mimics. Figure 1 shows the extracted features and a few samples of images with blood vessels and sulci removed. We exploited the elongated tubular structure of vessels/sulci to extract the following edge and orientation-based features:

**Figure 1:**
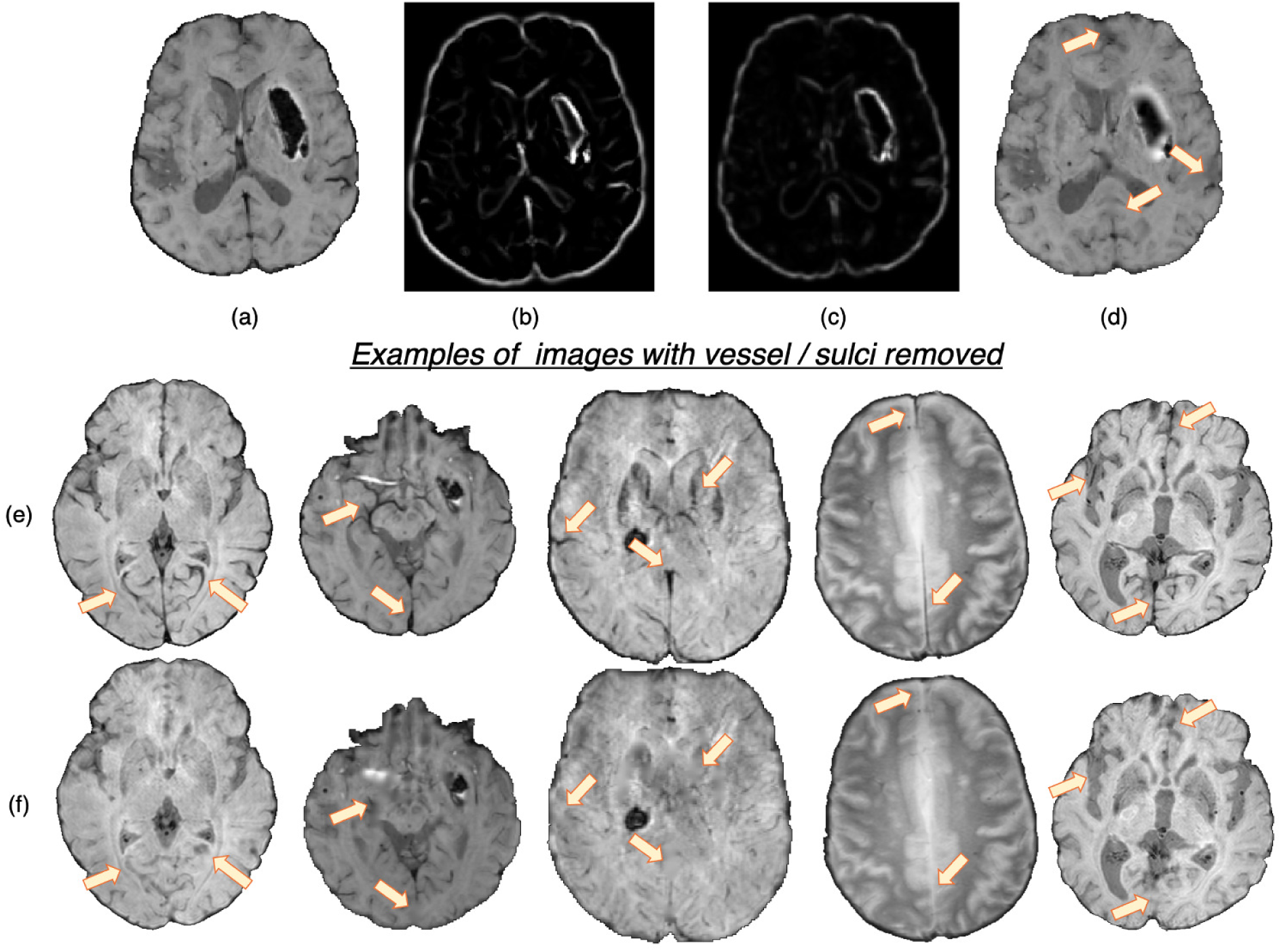
Features used for the vessel/sulci removal, along with a few samples of images with vessel/sulci removed. Top panel: (a) Input image, (b) Frangi filter output, (c) Structure tensor linearity measure output and (d) image with vessel / sulci removed. Note that the edges of the haemorrhages have also been smoothed out, aiding in false positive reduction. Bottom panel: A few instances of images shown in (e), along with their vessel removed results in (f). The arrows indicate the areas of noticeable vessel/sulci removal.

1. **Frangi filters:** Frangi filters (Frangi et al., 1998) use the multiscale second order local structure of the image and denote the degree of vessel-like/edge characteristics at each voxel. We extracted voxel-wise intensity values of Frangi filter outputs as features (figure 1b). While applying Frangi filters, we adjusted the parameters *β*_1_ and *β*_2_ (controlling the discrimination of lines from blob-like structures and the elimination of background noise respectively) to avoid detection of CMBs during vessel detection. Based on the results on the training data, we empirically set the values of *β*_1_ and *β*_2_ to 0.9 and 20.
2. **The eigenvalues of the structure tensor:** The structure tensor is the covariance matrix, at each voxel, consisting of partial derivatives of the gradients (Förstner, 1994). The eigenvalues *λ*_1_ and *λ*_2_ of this matrix indicate the edge strength. We considered the combination of the principal eigenvalue *λ*_1_ and the linearity measure *l* = |*λ*_1_ *− λ*_2_|*/*2 (figure 1c) as voxel-wise features.

We used K-means clustering, an unsupervised learning algorithm, with the above features to classify voxels into 2 classes (vessel vs background). The detected voxels with vessel-like structures and sulci are used as masks for inpainting. We filled the masked regions with intensity values similar to those in the non-masked neighbourhood (mean of the nearest 3 voxels) to remove linear dark structures from the input image.

### 3.2. Voxel-wise detection of initial CMB candidates

In the second step, as shown in figure 2, we extracted the following 7 features at each voxel on the vessel-removed image for the initial CMB candidate detection:

1. **Intensity transformations (3 features):** At each voxel, we extracted the intensity value (figure 2a). Additionally, to obtain contrast enhanced intensity features, we normalised the intensity values using standardisation (subtracting the mean intensity within the brain mask and dividing by standard deviation), extracted the exponential of the intensity (exp(p × intensity), where p = 1) (figure 2b) and applied ‘contrast limited adaptive histogram equalisation’ (CLAHE, Zuiderveld (1994), using *equalize adapthist* in *scikit-image* package) with a clip limit of 0.01 on the input image to obtain the CLAHE output at each voxel (figure 2c).
2. **Fast radial symmetry transform (FRST):** We performed FRST (Loy and Zelinsky, 2002). In FRST, at each voxel, the orientation (pointing towards or away from the voxel) and magnitude maps of gradients are calculated at a certain radius. These maps are then used to obtain a voxel-wise symmetry information within the radius. We used 4 different radii: 2, 3, 4 and 6 voxels, and calculated the mean value of all 4 outputs at each voxel as a feature (figure 2d).
3. **The eigenvalues of structure tensor:** We considered the principal eigenvalue *λ*_1_ (since this reduced the noise in background voxels) of the structure tensor at each voxel as a feature (figure 2e).
4. **Gaussian filter:** We smoothed the vessel removed image *I* with a Gaussian filter with *σ* = 1.5 voxels (empirically determined to roughly match the size of CMBs) to get a smoothed image *I*_*S*_. The difference *I*_*δ*_ = *I − I*_*S*_ removes the background and highlights the sharper objects and blobs, and hence can be interpreted as a ‘blobness’ measure (figure 2f).
5. **Laplacian of Gaussian filter (LoG):** Since CMBs have well-defined edges, we applied LoG (with *σ* = 1.5 voxels), a second order derivative filter for edge detection, on the vessel removed image and obtained the filtered output (figure 2g) at each voxel.

**Figure 2:**
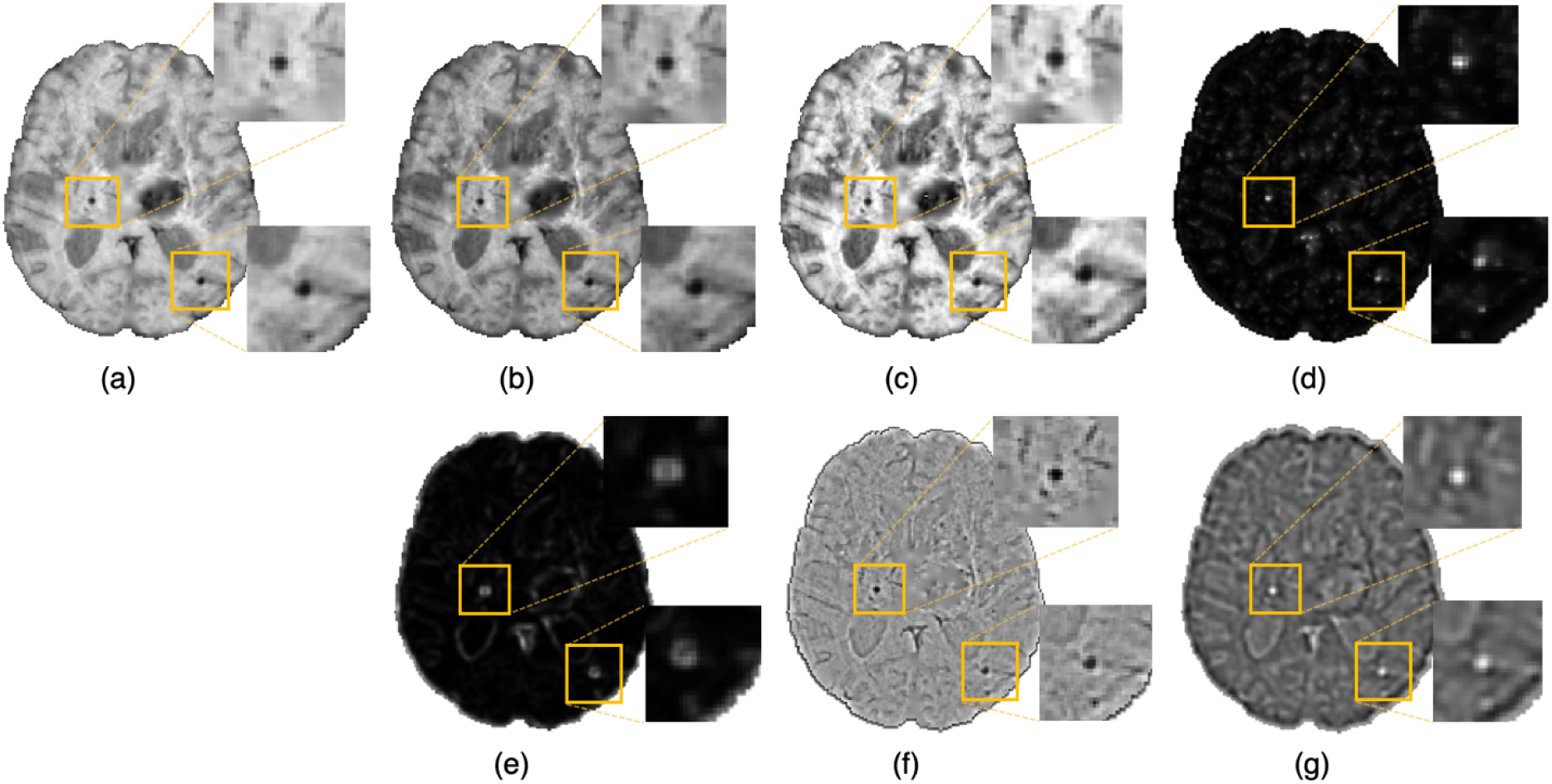
Features extracted for the voxel-wise CMB candidate detection. Images showing (a) Input intensity, (b) exponential transformed intensity, (c) CLAHE output, (d) fast radial symmetry transform (FRST) output, (e) structure tensor output, (f) Gaussian filtered output and (g) Laplacian of Gaussian (LoG) output. Inset figures show the magnified versions of the regions indicated in the boxes.

We normalised the above features individually by dividing by their maximum value across the current image and use the normalised features for training a support vector machine (SVM) classifier at the voxel-level, using voxel-wise manual segmentations available for the OXVASC and TICH2 datasets, to obtain a probability map *P*_*CMB*_. We then thresholded it at a global threshold *th*_*prob*_ of 0.8. The threshold value was determined empirically (using the training data from the OXVASC and TICH2 datasets), based on the cluster-wise performance metrics with respect to lesion-level manual segmentations, by varying *th*_*prob*_ within a range [0, 1] (for the details on the cluster-wise evaluation metrics used and the results, refer to the suppl. section 2).

### 3.3. Filtering of initial candidates using shape-based attributes

The small dark structures in the image other than CMBs (including noise and stray fragments of blood vessels) are detected as false positives (FPs) in the voxel-level classification. In this step, we used the following shape- and location-based object-level attributes for reducing FP as shown in figure 3:

1. **Volume** *V*_*c*_ of the candidate in mm^3^. Candidates with 5 mm^3^ *< V*_*c*_ *<*120 mm^3^ were selected as CMBs.
2. **Ellipticity** *ε*_*c*_: A measure of elongated nature of an object. The value ranges between [0,1] and a sphere has a value of 0. Candidates having *ε*_*c*_ *<* 0.2 were selected as CMBs.
3. **Solidity** *S*_*c*_: The ratio between volume of the candidate and its convex volume. The value ranges within [0,1] and an object with its volume equal to that of its convex hull has a value of 1; this criterion removes the curved fragments of blood vessels and intersection of vessels that survived the vessel removal step, since they would have low solidity. A lower threshold value of 0.6 was applied on *S*_*c*_ of candidates to be selected as CMBs.
4. **Diameter** *D*_*c*_ of the candidate in mm. Here, diameter is the distance between the endpoints of the longest line that can be drawn through the candidate. Candidates with diameters 2mm *< D*_*c*_ *<* 10mm were selected as CMBs.

**Figure 3:**
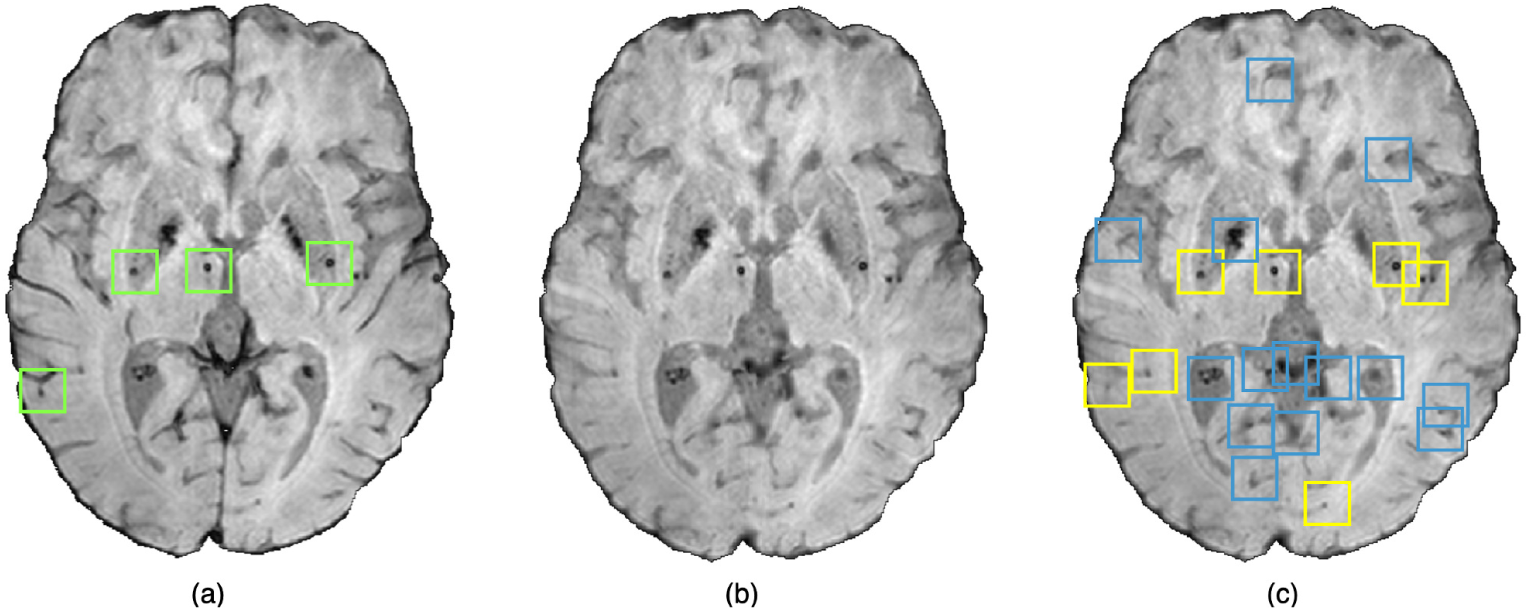
An instance of the shape-based filtering of initial candidates. (a) Input image with green boxes indicating manually segmented CMBs, (b) vessel removed image and (c) result of the shape-based filtering. In the output (c), blue boxes indicate initial candidates that were then rejected by the filtering stage and yellow boxes indicate the final CMB candidates that survived the filtering stage.

We considered a candidate as a CMB if all the above criteria were satisfied. The values for the above criteria were chosen empirically by a trial-and-error method based on the best subject-level performance on the training data from the OXVASC and TICH2 datasets using the metrics specified in section 3.5. The final cluster-wise performance values (providing lesion-level CMB detection performance) after the filtering step are provided in suppl. section 2.2.1.

#### Preselection criterion

Finally, a subject is classified as having CMBs if the count of the detected CMB candidates (after the filtering stage) exceeds an empirically set threshold value *Th*_*NCMB*_. For each dataset, we determined the *Th*_*NCMB*_ value by measuring the subject-level performance values (specified in section 3.5) at various thresholds within a pre-specified range, and choosing the threshold that provided the best set of performance values as *Th*_*NCMB*_ (refer to section 4.1). The range of threshold values was set to be slightly higher than the average number of CMBs in the OXVASC and TICH2 datasets since we had to allow for the presence of false positives (e.g. vessel fragments and sulci missed in the vessel removal stage).

### 3.4. Evaluation of the CMB candidate preselection pipeline

We evaluated the proposed pipeline by training and testing it using different datasets (with different modalities and acquisition characteristics) to study the effect of dataset characteristics on its performance. We performed the following experiments:

#### 3.4.1. Initial evaluation of the proposed preselection pipeline within datasets

We performed leave-one-out validation separately on the OXVASC and TICH2 datasets. We used T2*-GRE images from 40 subjects from the OXVASC dataset and, separately, SWI from 50 subjects from the TICH2 dataset. For the UKBB dataset, we performed 5-fold cross-validation on SWI images from 180 subjects with training/validation/test split of 70/10/20%. Note that, for the cross-validation on the UKBB dataset, we used the SVM model trained on the TICH2 dataset for the initial candidate detection (since voxel-wise manual segmentations were not available for the UKBB dataset). We determined the performance metric values (specified in section 3.5) at different settings of the threshold *Th*_*NCMB*_ in order to plot the ROC curve.

#### 3.4.2. Comparison of the proposed pipeline with algorithms using imaging/non-imaging factors on the UKBB dataset

In addition to the CMB lesion count extracted from imaging data, various non-imaging demographic/clinical factors have been associated with the incidence of CMBs, with age and BP being the common ones (Roob et al., 2000; Horita et al., 2003; Vernooij et al., 2008; Poels et al., 2010). We compared the following category of methods (including the proposed method) using imaging and/or non-imaging information:

1. **Method 1: demographic/clinical factors:** We considered factors such as age, diastolic BP and systolic BP separately by applying a range of thresholds to each factor to determine the baseline performance of each factor.
2. **Method 2: other non-imaging factors:** While age and BP are the most common factors, several other factors have also been associated with CMBs. Therefore, we considered other non-imaging factors such as smoking, white matter hyperintensity (WMH) volume and mean reaction time (MRT, linked with cognitive ability) in addition to age and BP. We used these 6 factors as features to train a SVM classifier (*SV M*_*NI*_) and a random forest classifier (*RF*_*NI*_). For *SV M*_*NI*_, we used a radial basis function kernel, with tolerance value of 1 × 10^*−*3^ and *E* of 0.1 (used *fitrsvm* in Matlab (2016b)). For *RF*_*NI*_, we used 150 trees, samples at leaf node = 5, mean squared error (MSE) as criteria for splitting and out-of-bag error score set to True in *TreeBagger* command in Matlab (for getting feature importance, see below). The above training hyperparameters were chosen using trial-and-error method on the validation data. We performed 5-fold cross-validation with the same training-validation-test split used in section 3.4.1. We also performed feature ranking to determine the importance of individual features using the out-of-bag (OOB) prediction^2^ error metric in *RF*_*NI*_.
3. **Method 3: determining CMB lesion count using imaging-based methods:** Similar to our proposed pipeline, we used imaging data (T2*-GRE from OXVASC and SWI from TICH2) and applied another baseline method, where we replaced the SVM-based CMB candidate detection step with a thresholding method to detect initial CMB candidates. We used the same preprocessing steps used in our pipeline (refer to section 2.1) and applied a threshold value at the lower 5^th^ percentile of the intensity value (determined empirically from the intensity histograms) and considered the voxels within the brain mask below the threshold as initial CMB candidates. We compared the 5-fold cross-validation results of our proposed method with the thresholding method.
4. **Method 4: non-imaging factors + CMB lesion count determined from imaging data:** We used a total of 7 features including the 6 factors used in method 2 and the CMB lesion count (determined using the proposed pipeline). Similar to method 2, we trained a SVM (*SV M*_*NI*+*I*_) and RF (*RF*_*NI*+*I*_) classifiers and evaluated them using 5-fold cross-validation. In addition, we determined the feature importance from OOB prediction error using *RF*_*NI*+*I*_. For training the RF and SVM classifiers, we used the same parameters used in method 2.

From all the above methods we determined the performance values (specified in section 3.5) at different thresholds for plotting the ROC curves.

#### 3.4.3. Evaluation of the generalisability of our method across datasets

We trained our pipeline (SVM classifier in the candidate detection step) on T2*-GRE images from 40 subjects of the OXVASC dataset (and performed hyper-parameter optimisation on the held-out data from the OXVASC dataset as specified in suppl. section 2). We later evaluated it on the SWI from 50 subjects of the TICH2 dataset. Similarly, we trained the pipeline on SWI from the TICH2 dataset and evaluated it on T2*-GRE images of the OXVASC dataset. Finally, we applied both OXVASC-trained and TICH2-trained pipelines on SWI from 180 subjects of the UKBB dataset. We could not train the SVM model on the UKBB dataset due to non-availability of voxel-wise manual labels and hence all training/testing combinations were not possible. For these experiments, we determined the performance of the preselection pipeline using the evaluation measures specified in section 3.5.

### 3.5. Performance evaluation metrics for CMB candidate subject preselection pipeline

For the CMB candidate subject preselection pipeline, we used the following measures for evaluation:

- **Subject-level true positive rate (TPR):** For a given dataset *D*, the subject-level TPR is the number of predicted TP CMB subjects (*S*_*TP*_) divided by the number of true CMB subjects, as given by,

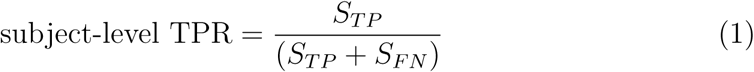

where *S*_*FN*_ is the number of false negative subjects.
- **Subject-level specificity:** For a given dataset *D*, the subject-level specificity is the number of predicted true negative subjects (*S*_*TN*_) divided by the number of non-CMB subjects, as given by,

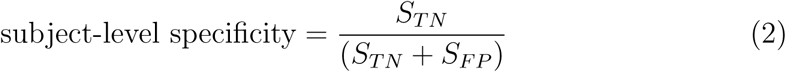

where *S*_*FP*_ is the number of false positive subjects.
- **Subject-level accuracy:** For a given dataset *D* consisting of *S*_*N*_ subjects, the subject-level accuracy is given by the number of correctly predicted CMB and non-CMB subjects (*S*_*TP*_ and *S*_*TN*_) divided by the total number of subjects,

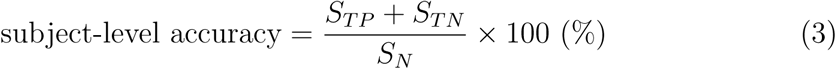

We plotted subject-level ROC curves using subject-level TPR and subject-level false positive rate (subject-level FPR = 1 - subject-level specificity) values.

Additionally, although it was not our main focus, we also determined various cluster-wise measures for obtaining an indicative evaluation of the lesion-level performance at CMB initial candidate detection step (refer to section 2 of the suppl. material for more details).

## 4. Results

### 4.1. Initial evaluation of the proposed preselection pipeline within datasets

Figure 4 shows the separate leave-one-out validation results for the OXVASC and the TICH2 datasets. The best set of performance values for each dataset was determined from the knee point on the ROC. From the ROC curves it can be seen that, on the OXVASC dataset, the pipeline achieves the best subject-level performance metric values: TPR = 0.96, specificity = 0.83 and accuracy = 0.90 at the threshold *Th*_*NCMB*_ = 30 CMBs (and TPR = 0.81, specificity = 0.87 and accuracy = 0.84 at the threshold *Th*_*NCMB*_ = 35 CMBs). Similarly, on the TICH2 dataset, the pipeline achieves the best subject-level performance values: TPR = 0.91, specificity = 0.81 and accuracy = 0.86 at the threshold *Th*_*NCMB*_ = 35 CMBs. On performing the 5-fold cross-validation on the UKBB dataset (ROC curve shown by black solid line in figure 5), the proposed pipeline achieved the best subject-level performance with a TPR = 0.91, specificity = 0.86 and accuracy = 0.89 at *Th*_*NCMB*_ = 35 CMBs.

**Figure 4:**
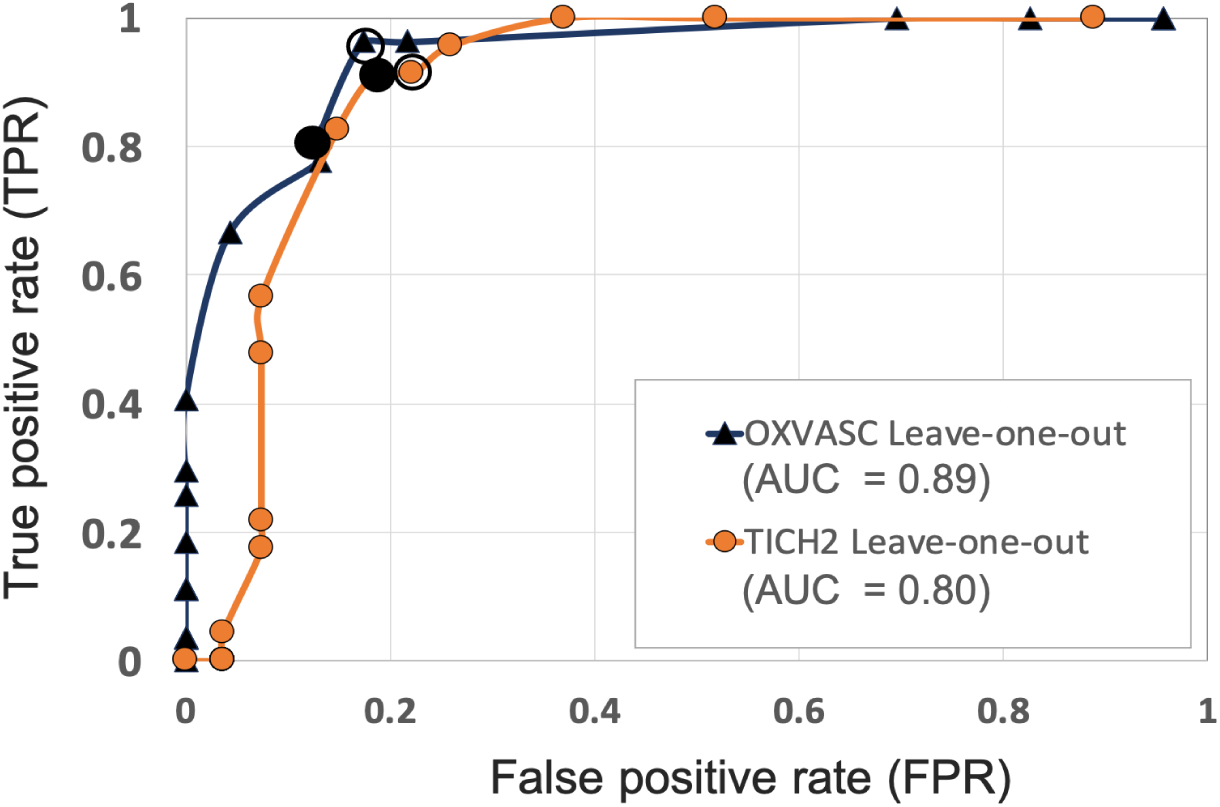
ROC curves for leave-one-out validations of the full preselection pipeline shown for the OXVASC (dark blue solid ▴) and the TICH2 (orange solid •) datasets plotted at threshold values from 10 to 80 CMBs in steps of 5 CMBs (AUC: area under the curve). The points on curves for the threshold value of 30 and 35 CMBs are indicated by hollow and filled black circular markers respectively. The performance values at the ‘knee point’ of curves were chosen as the best performance values for each dataset.

**Figure 5:**
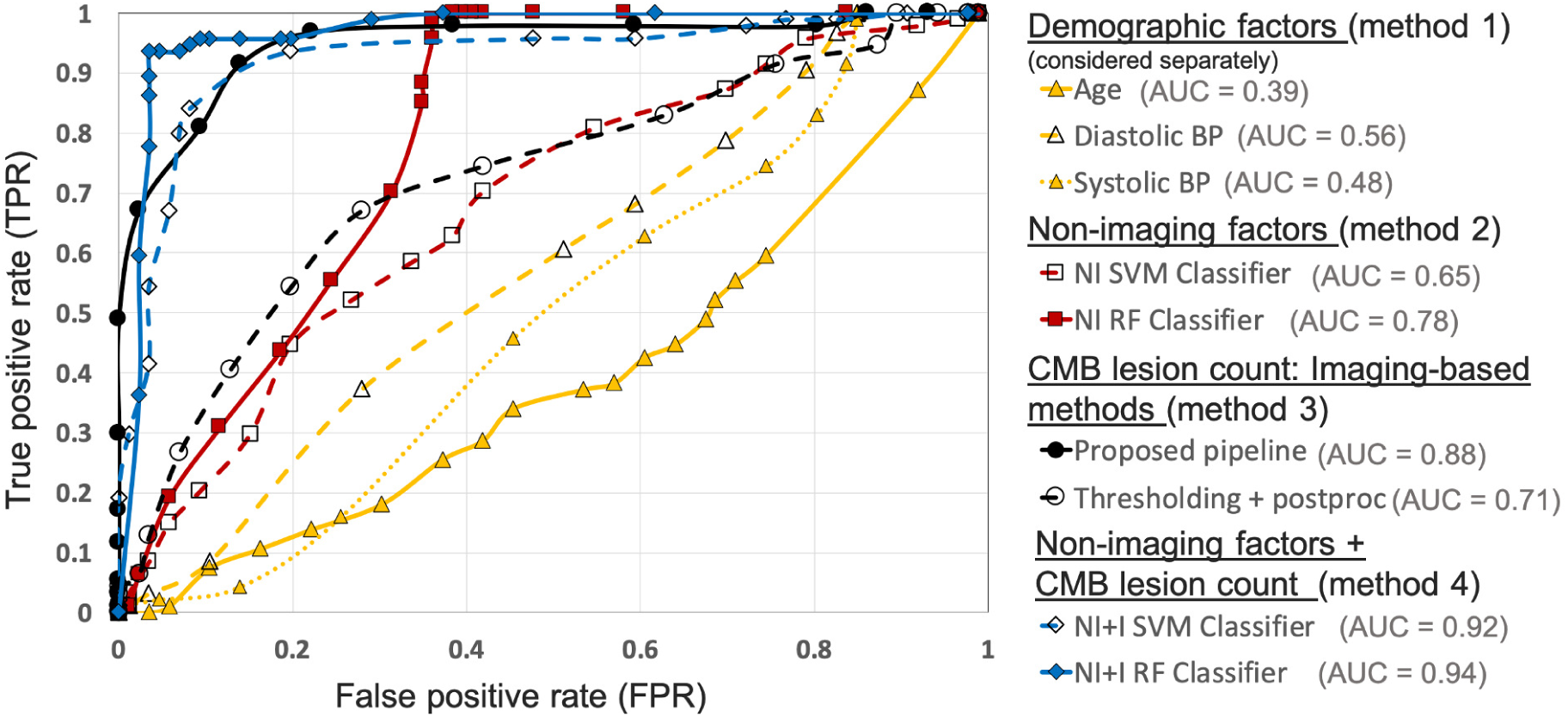
Results of the comparison of various methods for CMB candidate subject preselection on the UKBB dataset. ROC curves shown for thresholding on individual factors - age (yellow solid ▴, threshold range: 46.9 - 76.9 yrs, steps of 1.5 yrs), diastolic BP (yellow dashed ▴, threshold range: 50.9 - 118 mmHg, steps of 3.1 mmHg), systolic BP (yellow dotted △, threshold range: 106.4 - 202 mmHg, steps of 5.3 mmHg), classification based on non-imaging factors (NI) using an SVM classifier (red dashed □, threshold range: 0 - 1, steps of 0.05) and a RF classifier (red solid ▪, threshold range: 0 - 1, steps of 0.05), based on only the CMB lesion count obtained from the proposed pipeline (black solid •, threshold range: 0 - 90, steps of 5) and the thresholding method (black dashed ○, threshold range: 0 - 75, steps of 3), classification based on both non-imaging factors and CMB lesion count (NI+I) obtained from the proposed pipeline using an SVM classifier (blue dashed ◊, threshold range: 0 - 0.9, steps of 0.05) and a RF classifier (blue solid ♦, threshold range: 0 - 0.9, steps of 0.05) (AUC: area under the curve). The performance values at the ‘knee point’ of curves were chosen as the best performance values for each curve (for corresponding threshold values, refer to suppl. table 2).

### 4.2. Comparison of the proposed pipeline with algorithms using imaging/non-imaging factors on the UKBB dataset

Figure 5 shows the ROC curves for comparison of various methods used for preselecting CMB candidates from the UKBB dataset. For the best performance metrics from ROC curves for each method, refer to the suppl. section 3. Out of all the categories, using individual demographic/clinical factors (method 1) provided the worst performance. Among the individual factors, age provides the worst performance while the diastolic BP provided better performance, with subject-level TPR = 0.37, specificity = 0.72 and accuracy = 0.54 at a threshold of 93.4 mmHg. For classification based on non-imaging factors (method 2), the RF classifier provided better results compared to the SVM classifier. Using the RF classifier, we obtained subject-level TPR = 0.73, specificity = 0.74, accuracy = 0.74 at a threshold of 0.6. Among imaging-based methods (method 3) used to determine CMB lesion count, the proposed pipeline provided better results (subject-level TPR = 0.91, specificity = 0.86 and accuracy = 0.89) compared to the thresholding method, detecting more CMB lesions in the CMB subjects when compared to the non-CMB subjects, and hence better at identifying subjects containing CMBs. Of all the methods, classification using both non-imaging factors and CMB lesion count (method 4) provided the best performance, especially using the RF classifier, achieving subject-level TPR, specificity and accuracy of 0.95 at a threshold of 0.6.

Figure 6 shows the OOB feature importance of various features used in the *RF*_*NI*_ and *RF*_*NI*+*I*_ classifiers. For *RF*_*NI*_, WMH volume had the highest importance (imp = 2.1) followed by the BP values (diastolic BP imp = 0.41 and systolic BP imp = 0.42). For *RF*_*NI*+*I*_, CMB lesion count had the highest feature importance (imp = 2.8) followed by WMH volume (imp = 1.25) and diastolic BP (imp = 0.35). The higher feature importance of diastolic BP compared to age (imp = 0.25) also aligns well with our earlier comparison where diastolic BP provided the best performance among the individual factors. For both *RF*_*NI*_ and *RF*_*NI*+*I*_, smoking was the least important feature with imp = -0.08 and 0.01 respectively.

**Figure 6:**
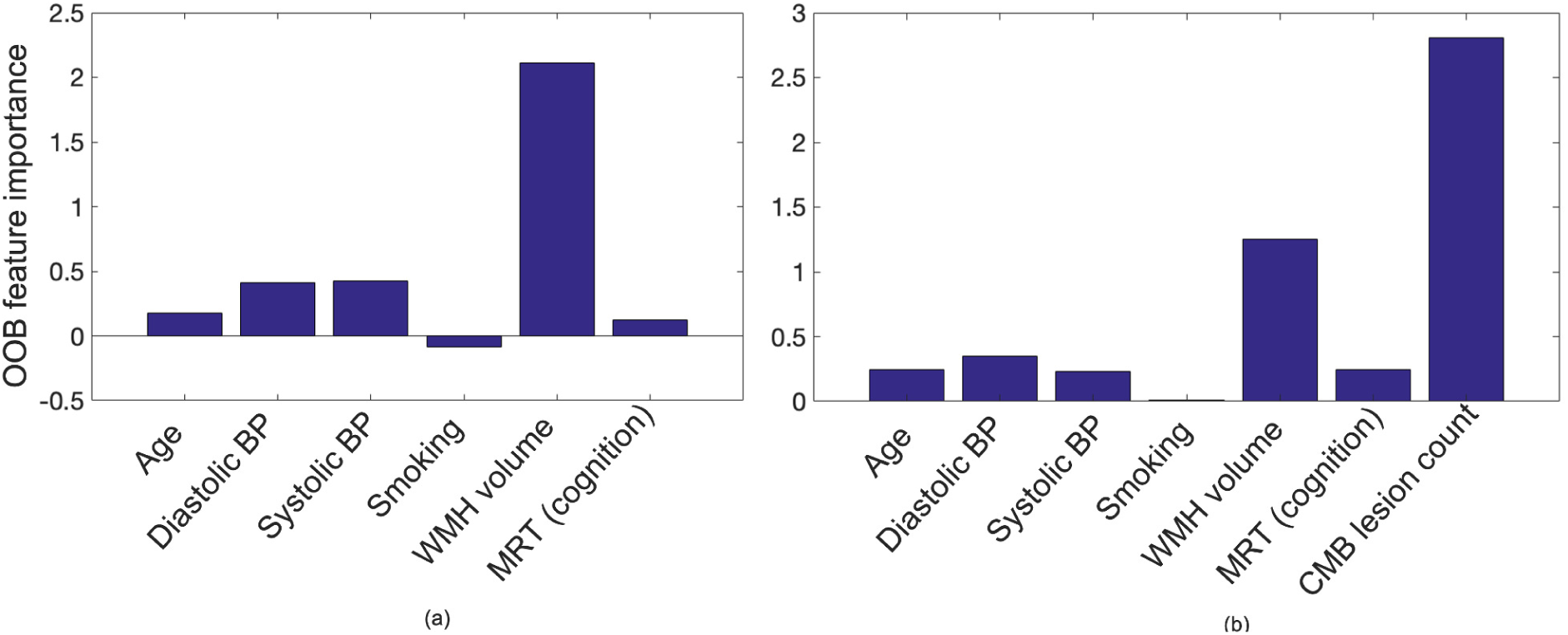
Out-of-bag feature importance values for (a) non-imaging features and (b) non-imaging features + CMB lesion count (obtained from the proposed pipeline) as used in the classification for CMB subject preselection.

### 4.3. Evaluation of the generalisability of the proposed pipeline across datasets

#### Pipeline trained on OXVASC and evaluated on the TICH2 data

Figure 7a shows the ROC curve for the prediction of CMB subjects at various values of the threshold *Th*_*NCMB*_ on the number of detected CMBs for individual subjects. We achieved the best performance with *Th*_*NCMB*_ set to 35 CMBs. Despite the presence of haemorrhagic lesions in all subjects, the pipeline gave a sensitivity of 0.90 and an accuracy of 0.81, with a subject-level specificity of 0.70. Figure 8 shows a few example cases of correct and incorrect subject-level detections on the TICH2 dataset (manually segmented CMBs are indicated with green boxes). From the figure, it can be seen that our algorithm correctly predicted the subjects with a high number of CMBs (figure 8a,b). Typically these subjects with high number of CMBs or no CMBs were detected better than subjects with very low number of CMBs (well under *Th*_*NCMB*_ value of 30 CMBs, as shown in 8e) or subjects having small haemorrhages (figure 8c, d).

**Figure 7:**
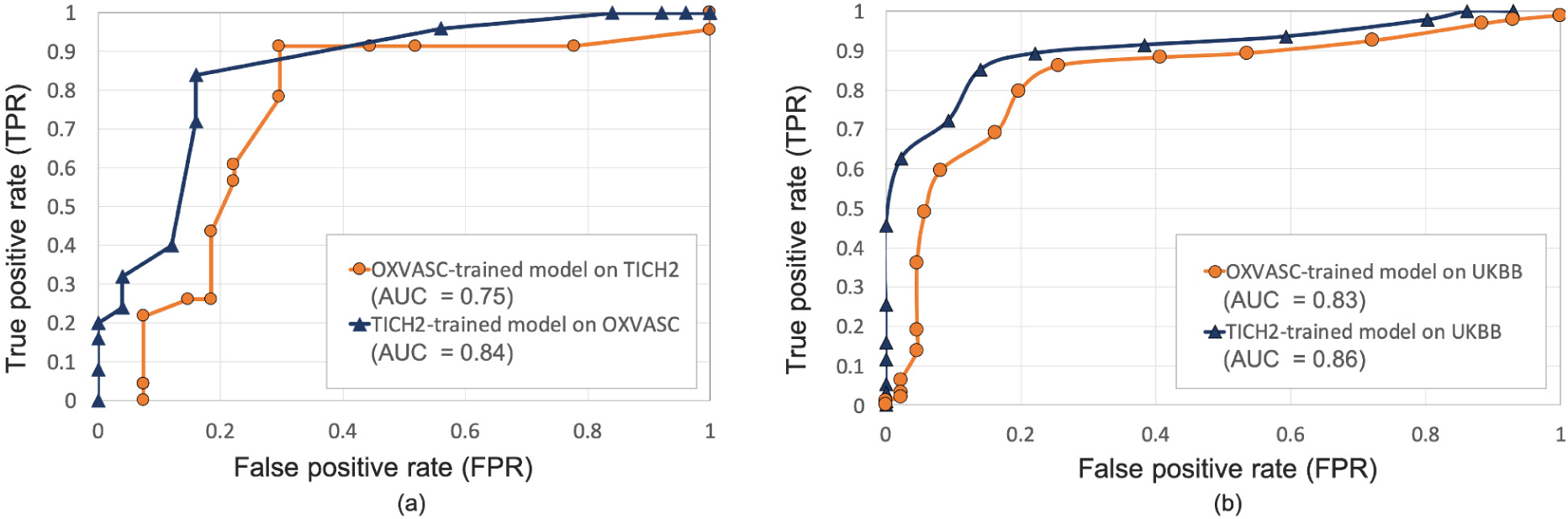
Results of the evaluation of model generalisability by training the pipeline on different dataset from the one it is evaluated on. ROC curves shown for (a) OXVASC-trained pipeline evaluated on the TICH2 dataset (orange) and TICH2-trained pipeline evaluated on the OXVASC dataset (dark blue), (b) OXVASC-trained and TICH2-trained pipelines evaluated on the UKBB dataset (orange and dark blue curves respectively) (AUC: area under the curve).

**Figure 8:**
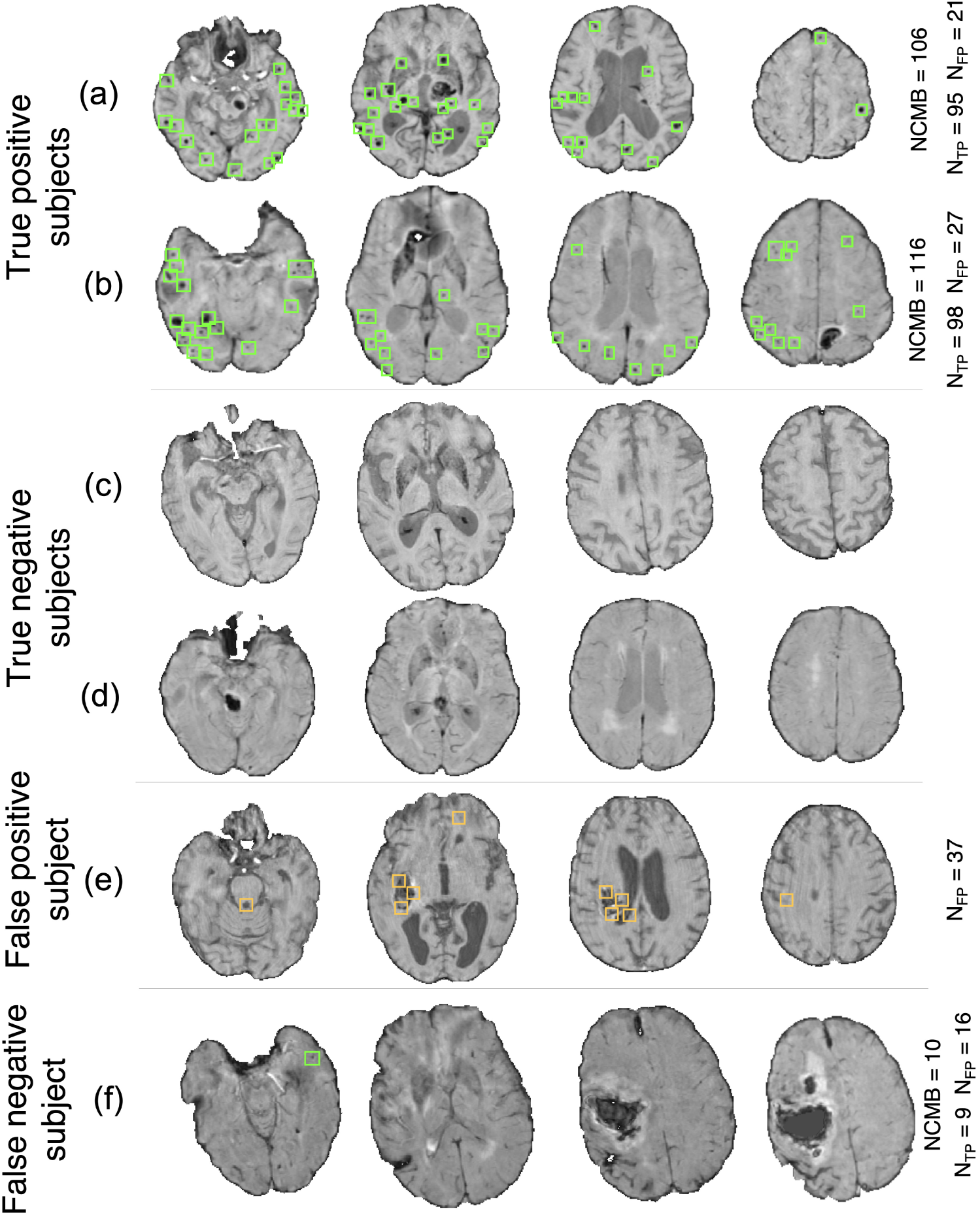
Example results from the CMB subject preselection pipeline trained on OXVASC data. (a) and (b) are true positive CMB subjects, (c) and (d) are true negative non-CMB subjects, (e) is a false positive prediction of non-CMB subject and (f) is a false negative prediction of a CMB subject. The green and orange boxes indicate the manually segmented CMBs and false positives respectively. The true CMB count *NCMB* are provided, along with the number of true positives (*N*_*TP*_) and the false positives (*N*_*FP*_).

#### Pipeline trained on TICH2 and evaluated on the OXVASC data

Figure 7a shows the ROC curve for CMB candidate subject preselection with the TICH2-trained model on the OXVASC dataset. As in the previous case, the model achieved the best performance for a threshold value *Th*_*NCMB*_ of 35 CMBs. The model provided a subject-level TPR of 0.83, a subject-level specificity of 0.84 and a subject-level accuracy of 0.83. The number of subjects with high CMB counts (*>* 50 CMBs) in the training dataset (TICH2) is higher than that in OXVASC. Hence, the TICH2-trained model was more specific in detecting CMB subjects on the OXVASC dataset, achieving a higher specificity of 0.84. We have shown a few examples of subject-level detections in figure 9. As in the previous case, subjects with high CMB counts (figure 9a and b) were correctly predicted as CMB subjects.

**Figure 9:**
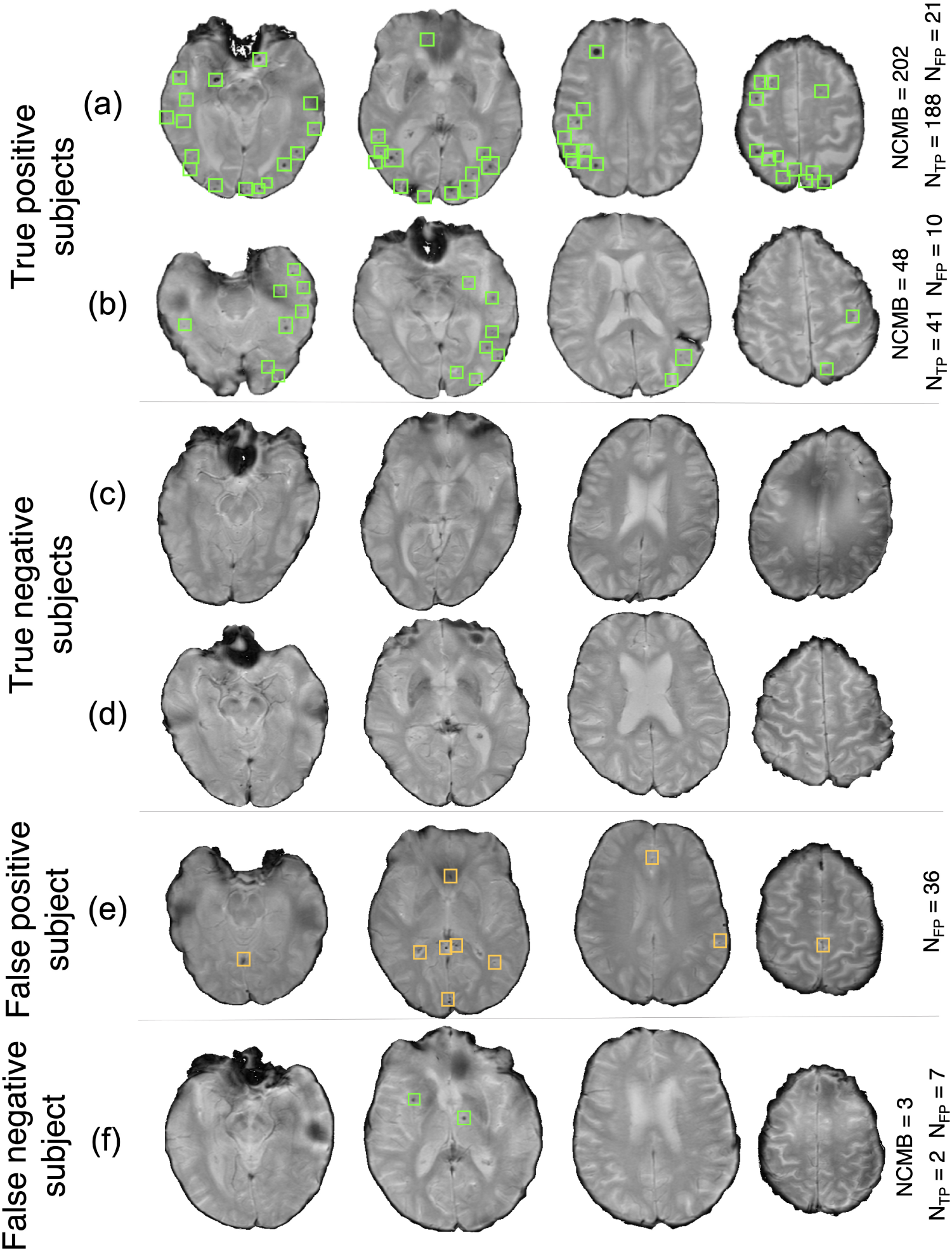
Example results from the CMB subject preselection pipeline trained on TICH2 data. (a) and (b) are true positive CMB subjects, (c) and (d) are true negative non-CMB subjects, (e) is a false positive prediction of non-CMB subject and (f) is a false negative prediction of CMB subject. The green and orange boxes indicate the manually segmented CMBs and false positives respectively. The trueCMB count *NCMB* are provided, along with the number of true positives (*N*_*TP*_) and the false positives (*N*_*FP*_).

#### OXVASC-trained and TICH2-trained pipelines on the UKBB dataset

We applied both the OXVASC-trained and the TICH2-trained models to the UKBB dataset. The model trained on the TICH2 dataset provided better performance when compared to the OXVASC-trained model. For the OXVASC-trained model, the pipeline achieved subject-level TPR = 0.80, specificity = 0.80 and accuracy = 0.80 using a threshold of 30 CMBs. For the TICH2-trained model, the pipeline achieved the subject-level TPR = 0.85, specificity = 0.86 and accuracy = 0.85 using a threshold of 35 CMBs. We could not perform the lesion-level analysis on the UKBB dataset (as we did for the OXVASC and TICH2 datasets), since we had only subject-level manual labels.

## 5. Discussion and conclusions

In this work, we proposed a fully automated pipeline, which is computationally light and takes into account various intensity, shape and anatomy-based characteristics of CMBs, for preselecting CMB candidate subjects from large datasets. We compared various methods involving non-imaging demographic/clinical factors and CMB lesion count from the imaging data, including our proposed pipeline, on a subset of the UKBB dataset for which we had subject-level manual labels regarding the presence of CMBs. We finally applied our pipeline on different datasets for training and evaluation to validate its generalisability with respect to variations in intensity, acquisition protocols and pathological conditions. Our pipeline provided subject-level accuracy *>*85% during initial validation on each individual dataset. On applying our pipeline on various datasets, we observed good generalisability across datasets with subject-level accuracy *>*85% when trained on SWI and applied to SWI/T2*-GRE images, and *>*80% when trained on T2*-GRE images and applied to SWI.

During the initial within-dataset evaluation, the pipeline provided better subject-level results on the OXVASC and UKBB datasets when compared with the results from the TICH2 dataset. This could be due to the fact that pipeline detected more false positive lesions along the edges of intracerebral haemorrhages in the TICH2 dataset. In addition to the presence of haemorrhages, the increased amount of FPs could be related to the use of SWI. In fact, the best threshold on the CMB lesion count (corresponding to the ‘knee point’ on the ROC curve) for subject-level is preselection is higher for datasets with SWI (the threshold is 35 CMBs for both the UKBB and TICH2 datasets) when compared to the OXVASC dataset with T2*-GRE images (where the threshold is 30 CMBs). This might be because SWI improves not only the contrast of CMBs but also the mimics that could lead to more spurious CMB detections and hence a higher threshold is needed on the CMB lesion count. However, while we chose thresholds that gave the best performance on the datasets, we cannot exclude the possibility of subjects with low number of true CMBs (with low number of false positives) being incorrectly classified as non-CMB subjects.

On comparing various methods involving non-imaging and CMB lesion counts from the imaging data, we observed that applying thresholds on the individual demographic factors provided the worst results. This shows that even though factors such as age and hypertension are commonly associated with CMBs, they are not sufficient by themselves to preselect CMB candidate subjects in a given population. This further supports the need for pipelines such as ours that can also use the imaging data for subject-level preselection, especially in large datasets. Interestingly, among age and BP, the latter provides better preselection (and also has higher feature importance in the RF classifier), despite age being reported as the most commonly used factor (Vernooij et al., 2008; Poels et al., 2010; Takashima et al., 2011). Similarly, among the features used in the RF classifiers (both *RF*_*NI*_ and *RF*_*NI*+*I*_), smoking is the least important feature, even though it has been reported as one of the risk factors for CMBs in various population-level studies (Tsushima et al., 2002; Poels et al., 2010). However, more experimentation would be required on a larger sample size to further establish the relationship between age/smoking status and occurrence of CMBs.

The proposed method provided better results than the thresholding method, highlighting the utility of shape-based features, in addition to intensity. The best results were obtained for the classifiers using a combination of non-imaging factors and CMB lesion counts, however, with CMB lesion count being the most important feature. The high feature importance of CMB lesion count shows that the proposed pipeline determines a lesion count that is highly useful, despite the number of false positive lesions. Also, the best performance of the combination of features and lesion counts shows that the non-imaging features could be used to improve the results of our proposed pipeline for CMB candidate subject preselection in large datasets, when manual segmentation is unavailable. While the preselected subjects could be used for manual labelling for research purposes, from a clinical point of view the pipeline could be used to flag images as likely to contain CMBs. This could be further used in determining a preliminary CMB or small vessel disease (SVD) score as done in Staals et al. (2014).

On evaluating the generalisability of the proposed pipeline, the performance when the training and testing datasets were acquired with different modalities was slightly lower than the initial cross-validation performance values. Our preselection pipeline provided results with similar accuracy for different training data, with a slightly better specificity on the OXVASC dataset using TICH2-trained model when compared to the TICH2 dataset using OXVASC-trained model, while providing a lower subject-level TPR. The prediction of CMB candidate subjects was more precise in the OXVASC dataset likely due to the fact that the pipeline was trained using features extracted from SWI that is highly sensitive to CMBs and susceptibility artefacts. At this point, it is worth noting that there are pros and cons with using T2*-weighted GRE or SWI sequences. In the case of T2*-GRE images (the OXVASC dataset), the contrast between CMBs and background is lower than in SWI (and some CMBs might not be visible at all on T2*-GRE images) and hence the number of detected false positives was low, providing high specificity (85%, when compared to the specificity of 70% in the TICH2 dataset) but low cluster-wise TPR (83% compared to the TPR of 90% in the TICH2 dataset). This shows that SWI sequences (the TICH2 dataset) are highly sensitive to CMBs and hence result in detection of more CMBs, albeit with increased FPs as well. Therefore, choosing training datasets containing a similar modality (especially those having single modalities) to train the pipeline could help in achieving more accurate detection when applied to the large datasets. In fact, on applying OXVASC-trained and TICH2-trained pipelines to the UKBB dataset, the TICH2-trained pipeline provided better results since it was trained on SWI, the modality used in the UKBB dataset. However, it is worth noting that, despite the difference in population and CMB prevalence across the datasets used in this study, our pipeline showed comparable performance, even when trained on a dataset with very different prevalence (e.g. OXVASC, a stroke population, and UKBB, a prospective epidemiological study).

Concluding, we proposed a learning-based method for subject-level preselection of CMB candidate subjects in large datasets. The preselected subjects could then be manually segmented and used for further analysis and characterisation of CMBs. Our method provided accurate preselection of CMB candidate subjects on various datasets consisting of T2*-GRE and SWI images with subject-level TPR, specificity and accuracy values *>*90%, *>*80% and *>*85% respectively. Also, our method is computationally efficient, and provided greater performance when compared to other methods using non-imaging factors and thresholding methods for obtaining CMB lesion counts from the imaging data. Our pipeline shows good generalisability across across various datasets providing subject-level accuracy *>*80%, and even *>*85% when applied to datasets with the same modality. The future direction of this work would be to improve the detection of CMBs at the lesion-level using deep learning and increase the model generalisability across different modalities. Also, another potential avenue of research would be to provide automated ratings of CMBs using their size and count to provide information that is consistent with clinical rating scales such as MARS scale.

## Data Availability

Requests for data from the OXVASC Study will be considered by P.M.R. in line with data protection laws. The TICH2 MRI substudy data can be shared with bonafide researchers and research groups on written request to the substudy PI Prof Rob Dineen. Proposals will be assessed by the PI with advice from the trial TICH2 Steering Committee if required and a Data Transfer Agreement will established before any data are shared. The UK Biobank datasets are available to researchers through an open application via https://www.ukbiobank.ac.uk/register-apply/.

## Acknowledgements

The data used in this work was obtained from UK Biobank under Data Access Application (8107, 36509, 43822). We are grateful to UK Biobank for making the resource data available, and are extremely grateful to all UK Biobank study participants, who generously donated their time to make this resource possible. We thank the Cambridge University Hospital BRC. We thank all the OXVASC study and TICH2 MRI substudy participants. For the OXVASC dataset, we acknowledge the use of the facilities of the Acute Vascular Imaging Centre, Oxford. We also thank Dr. Chiara Vincenzi and Dr. Francesco Carletti for their help on generating the manual masks used in our experiments.

## Footnotes

2 In RF, OOB predictions are those obtained on the samples that were left out while randomly choosing the training subsets for individual trees during bootstrap aggregating (bagging). We considered the increase in MSE while permuting the OOB observations across each feature, averaged over all trees in the ensemble and divided by the standard deviation obtained over the trees. Hence, larger increase in MSE indicates more important features.

