## Supplementary material for "Automated Detection of Candidate Subjects with Cerebral Microbleeds using Machine Learning"

#### 1 Cerebral microbleed (CMB) mimics and their characteristics

| CMB mimics | Description and characteristics |
|---|---|
| Cavernous malformations | Visible on T2-weighted images (subtypes I - IV). Leakage of blood from the vessels at various stages of degradation [1], with subtype II lesions appearing with popcorn-like structure (looks similar to ringing artefacts found on T2*-weighted images). Subtype - IV lesions are punctate and are more difficult to differentiate from CMBs [2]. |
| Haemorrhagic micrometastases | Metastatic melanoma often can bleed and appear hypointense on T2*-weighted images since melanin is paramagnetic, but has concomitant hyperintensities on T1-weighted images surrounded by oedema. Note that other than metastatic melanoma, other haemorrhagic metastases are possible. However, small non-oedemous lesions are difficult to distinguish from CMBs. |
| Diffuse axonal injury | Concomitant presence of abnormalities (e.g. skull fracture and contusions) [2, 3]. The clinical history of subjects is recommended for distinguishing from CMBs. |
| Small haemorrhages near infarction and ICH areas | Visible on T2-weighted, T2*-weighted, FLAIR, DWI sequences. Sometimes, they differ from CMBs in size. Would be easier to use infarctions as exclusion biomarkers. |
| Flow voids | Do not show blooming effect. They have linear/ curvilinear tubular structures extending through contiguous slices, usually appear in the cortical regions and are visible on T2-weighted images [2]. |
| Calcifications | They are usually found in the basal ganglia. However, calcifications could also occur in choroid plexuses and the pineal gland. They appear as hyperintense blobs on CT images [4]. |
| Partial volume artefacts (air-bone interfaces) | Do not show blooming effects. Particularly seen in frontal and temporal lobes (orbit and mastoid bones) and at the edges of the cerebellum. Distinguishable from CMBs using location priors and observation of contiguous slices. |

Table S1: CMB mimics and their distinguishing characteristics [1, 2].

### 2 Effect of threshold value on obtaining CMB candidates

In the CMB initial candidate detection step (section 3.2 in the main manuscript), we obtained the voxel-wise probability values from the SVM classifier in the initial probability map $P_{CMB}$. We had to apply a threshold $th_{prob}$ to this probability map to get binary maps defining the CMB initial candidates that could be passed to the next filtering step.

In order to determine a suitable $th_{prob}$ value for binarising $P_{CMB}$ map, we applied various threshold values from 0 to 1, increasing in steps of 0.1. For this experiment, we trained the SVM model on the OXVASC dataset and evaluated it on the TICH2 data and vice versa, since evaluating across datasets would provide a more robust threshold value. For each threshold value, we determined the cluster-wise evaluation metrics (specified below in section 2.1), and selected the threshold that provided the best set of cluster-wise performance metrics as the final $th_{prob}$ value.

#### 2.1 Cluster-wise evaluation metrics for selecting the optimal threshold $th_{prob}$

We determined the following cluster-wise metrics for the evaluating the lesion-level performance at the CMB initial candidate detection step:

- **Cluster-level TPR:** The number of true positive CMBs divided by the total number of true CMBs as given by,

$$\text{cluster-wise TPR} = \frac{TP_{clus}}{(TP_{clus} + FN_{clus})} \tag{1}$$

  where $TP_{clus}$ and $FN_{clus}$ are true positive and false negative CMBs respectively.

- **Average number of false positive clusters per subject (FPavg):** For a given dataset $D$, FPavg is defined as the ratio of the total number of detected FPs to the number of subjects (or images) in the dataset, as given by,

$$FPavg = \frac{\text{Total number of FPs}}{\text{Number of subjects in } D} \tag{2}$$

We used 26-connectivity to determine the clusters for obtaining above metric values. Also, in the above definitions, we considered a cluster as a true positive cluster if there is it overlaps with a ground truth cluster by at least one voxel. And false positive clusters are the ones that has no overlap with a ground truth cluster. We used TPR and FPavg values for plotting a free-response ROC (FROC) curve, which is a plot of TPR versus the average number of false positives per image/subject.

#### 2.2 Results

Figures S1a and b show FROC curves for the TICH2 and OXVASC datasets used to determine the threshold value $th_{prob}$ for the voxel-wise initial CMB candidate detection of the OXVASC-trained and TICH2-trained pipelines respectively.

Since we will be applying, at a later stage, the shape-based filtering step (step 3) to further reduce false positives, we prioritised achieving high cluster-wise TPR in this initial CMB candidate detection (step 2). For both the cases, we achieved the best performance at $th_{prob}$ value of 0.8 with FPavg values closer to or less than 200 CMBs/subject. The SVM classifier trained on the OXVASC dataset achieved a cluster-wise TPR of 0.906 with FPavg of 210.6 on the TICH2 dataset, while the SVM trained on the TICH2 dataset cluster-wise TPR of 0.86 with FPavg of 178.7 on the OXVASC dataset.

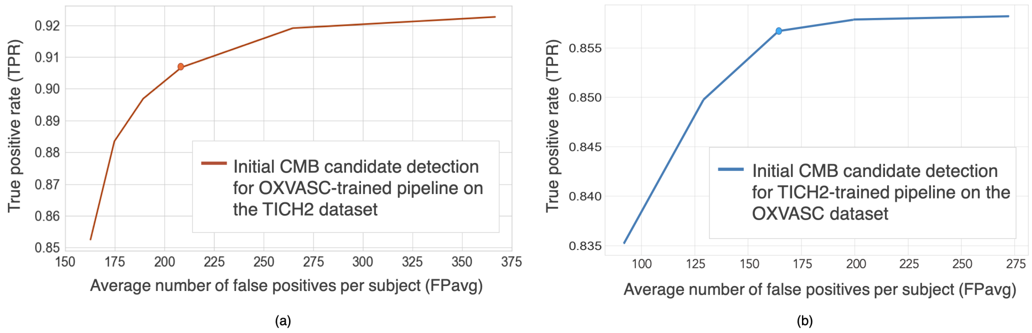


Figure S1: Free-response ROC (FROC) curves for initial CMB candidate detection for (a) OXVASC-trained pipeline evaluated on the TICH2 dataset and (b) TICH2-trained pipeline evaluated on the OXVASC dataset. The circular markers indicate the points at which we achieve the best compromise between TPR and FPs (the point that corresponded to the highest TPR having FPavg $\approx$ 200 FPs/subject.

##### 2.2.1 Cluster-wise results after filtering initial CMB candidates

For the pipeline trained on the OXVASC data and evaluated on the TICH2 data, the pipeline achieved a cluster-wise TPR of 0.88 with FPavg of 20.4 after he final candidate filtering stage. Similarly for the pipeline trained on the TICH2 data and evaluated on the OXVASC data, the pipeline achieved a cluster-wise TPR of 0.85 with FPavg of 12.8 after the final candidate filtering stage.

#### 3 Performance metrics for comparison of imaging and non-imaging based methods on the UKBB dataset

Table S2 shows the performance metrics for comparison of imaging and non-imaging based methods on the UKBB dataset along with the threshold values.

Table S2: Best performance values determined from the ROC curves for methods based on imaging (I) and non-imaging (NI) data, using the UKBB as the evaluation dataset (section 4.2). Best results in each category highlighted in bold. Sub. TPR: subject-level true positive rate (TPR), Sub. Spec: subject-level specificity, Sub. Acc: subject-level accuracy. For the final category (NI+I), the CMB lesion counts were determined by the proposed pipeline.

| **Methods** | **Sub. TPR** | **Sub. Spec** | **Sub. Acc** | **Threshold** |
|---|---|---|---|---|
| **Demographic/clinical factors considered individually** | | | | |
| Age | 0.34 | 0.55 | 0.44 | 62.7 yrs |
| **Diastolic BP** | 0.37 | **0.72** | **0.54** | 93.2 mmHg |
| Systolic BP | **0.46** | 0.55 | 0.50 | 148.9 mmHg |
| **Classification using non-imaging factors** | | | | |
| NI SVM classifier | **0.74** | 0.58 | 0.67 | 0.5 |
| **NI RF classifier** | 0.73 | **0.74** | **0.74** | 0.6 |
| **CMB lesion count: Imaging-based methods** | | | | |
| **Proposed pipeline** | **0.91** | **0.86** | **0.89** | 35 CMBs |
| Thresholding + postproc | 0.67 | 0.72 | 0.69 | 56 CMBs |
| **Classification using non-imaging factors + CMB lesion count** | | | | |
| NI+I SVM classifier | 0.82 | 0.89 | 0.85 | 0.5 |
| **NI+I RF classifier** | **0.95** | **0.95** | **0.95** | 0.6 |
